# Cumulative local recurrence rate is a misleading and non-representative outcome measure for early breast cancer trials

**DOI:** 10.1101/2024.09.11.24313382

**Authors:** Jayant S Vaidya, Max Bulsara, Uma J Vaidya, David Morgan, Michael Douek, Marcelle Bernstein, Chris Brew-Graves, Norman R Williams, Jeffrey S Tobias

**Affiliations:** Division of Surgery and interventional Science, University College London; Department of Biostatistics, University of Notre Dame; Imperial College Healthcare NHS Trust; Department of Radiation Oncology, Nottingham Breast Cancer Research Centre, University of Nottingham; Nuffield Department of Surgery, University of Oxford; Patient Advocate; Department of Imaging, University College London; Department of Radiation Oncology, University College London Hospitals

## Abstract

In many breast cancer radiotherapy trials, the results are presented in the form of cumulative incidence rates of local recurrence or Kaplan-Meier plots, in which deaths are censored. Censoring - using patients’ length of follow up until the point when they had last been seen alive – is included in the statistical model, under the correct assumption that they will continue to have a risk of developing a local recurrence. Censoring should be non-informative and balanced. However, if shorter follow up is unbalanced between treatments, or if shorter follow up is due to death (from whatever cause), these assumptions and therefore the model is no longer valid. It is therefore ambiguous to statistically ignore deaths when reporting local recurrence, by censoring them. We illustrate, with examples from randomised trials, why and how such graphs cannot give patients and clinicians a clear indication of the effects of treatments or prognosis. For instance, in one of these examples, 60% of patients were alive at 10 years, so those alive without a local recurrence should inevitably be lower than 60%, rather than the 90% estimated using the above method. The simple way to avoid this error is to turn the analysis on its head, by reporting chances of success rather than failure, by reporting the probability of being free of local recurrence (i.e. both death and local recurrence are events). This estimate truly represents what really happens to patients in terms of local control and the relative effectiveness of treatment(s) comprehensively. It also conforms with the recommendations of ICH-GCP, European (DATECAN) and American (STEEP) guidelines.

## Introduction

Many trials comparing long-term results of local treatments for breast cancer, especially radiotherapy trials, have presented their results as cumulative incidence of local recurrence (LR), or Kaplan-Meier plots, in which deaths are censored. Here we show that such graphs do not enable patients and clinicians to have a clear understanding of their prognosis. This point has been made previously^1-3^ but has not prevented the improper use of the Kaplan Meier model for estimating outcomes, when not all patients have the same duration of follow up and when some patients would have died during follow up^4^.

We illustrate our argument with examples from randomised clinical trials comparing two treatments for breast cancer^5-10^. We show why and how such graphs can be misleading. This conceptual recognition can promote improved presentation of trial data^11 12^.

While plotting these graphs^6-10 13^, considering only *local recurrences* as events would work well if everyone’s follow-up was the same and no one died. However, this of course is never the case because a) patients are never recruited all at the same instant in any trial, and b) some patients sadly also die (from any cause, not just breast cancer deaths) during follow up.

Censoring is a well-established solution for the first problem - not all patients will have the same length of follow up because they are not recruited at the same instant. For example, consider a trial which recruited from the year 2013 to 2022 and which is being analyzed in 2025. Even if all patients were last seen in 2025, the very first in the cohort would have 12 years of follow up, and the final one would have a maximum of 3 years of follow up. So, patients who are known to be alive at the time when they were last seen are ‘censored’ at the time of the last follow up. Censored patient data (length of follow up) until the point when they are last seen and ***were known to be alive*** at that time, are included in the analysis *with the assumption that they will continue to have a risk of having local recurrence beyond this point*.

However, for those trial participants who are known to have died, this assumption is, of course, no longer true. Therefore, censoring is not a solution for the second problem. Furthermore, for the model to provide accurate estimates in treatment trials, censoring must be independent of the treatment allocation and non-informative, so treatments should not differentially impact on patients being censored because of death. In a model in which patients are censored if they die, the assumption is also made that deaths are unrelated to the intervention or disease process. However, cancer treatments (systemic chemotherapy and external beam radiotherapy) can themselves have serious adverse events including death, and local or distant recurrence are also known to raise the risk of death. It is therefore ambiguous to statistically exclude all patients who died when reporting local recurrence.

It is in the quest for estimating “pure” local recurrence, with the mistaken belief that treatments have only local effects, that in the following examples, patients who died have been censored while plotting these graphs. Even if treatments had only a local effect, reiterating the point made above, such censoring at the time of death is inappropriate as these patients would be wrongly assumed to be at risk of local recurrence even after they have died.

The anomaly becomes particularly obvious for longer term results, as many more patients will die during the longer follow up. The results are further skewed when mortality is independently affected by treatment, and particularly when the direction of this effect is opposite to its effect on local recurrence. For example, if a treatment A vs B decreases the risk of local recurrence from 2% to 1% (‘*halving’*) but increases deaths from 4% to 5% (‘*only’* a 25% relative increase), it could be recommended as a better treatment to avoid local recurrence, when in reality the local control – the chance of being alive without local recurrence – may be the same (94%) in the two groups.

It is interesting that local recurrence appears to be almost unique amongst reported cancer outcomes, where an alleged *cumulative incidence* is reported in this way. For distant relapse, the outcome measure of ***disease-free survival*** is normally used, in which both relapse and death are counted as events, which results in the correct representation of reality, and provides the figure that represents the chance for a patient to be free of disease. We show that local recurrence free survival graphs better inform patients of long-term outcomes as they truly represent what they can expect to happen to them should they chose the treatment(s). Also, local recurrence has been regarded as a surrogate marker for survival and therefore cannot be analysed in isolation.

## Method

Here we reproduce graphic data from six well-known and influential papers. We do this to illustrate how a statistical method that censors deaths produces unrealistic results. In contrast, we demonstrate how to avoid this basic error using figures from two other recently published papers.

For each of these, we have placed side-by-side figures representing the outcomes of overall survival and local recurrence. We then illustrate both the percentage of patients alive, and also those living without local recurrence at a long-term follow up point (e.g., 10 years). We then assess whether these two estimates are consistent with each other and whether the latter can be truly representative of what really happens to our patients, and whether it represents a true long-term comparison between the two treatments for a patient or clinician contemplating the treatments at the outset.

## Results

The following examples illustrate how censoring the patients who have died leads to results that are self-contradictory, and do not give clinicians or patients the realistic picture of outcomes or comparison between outcomes of treatments.

**Example 1:**
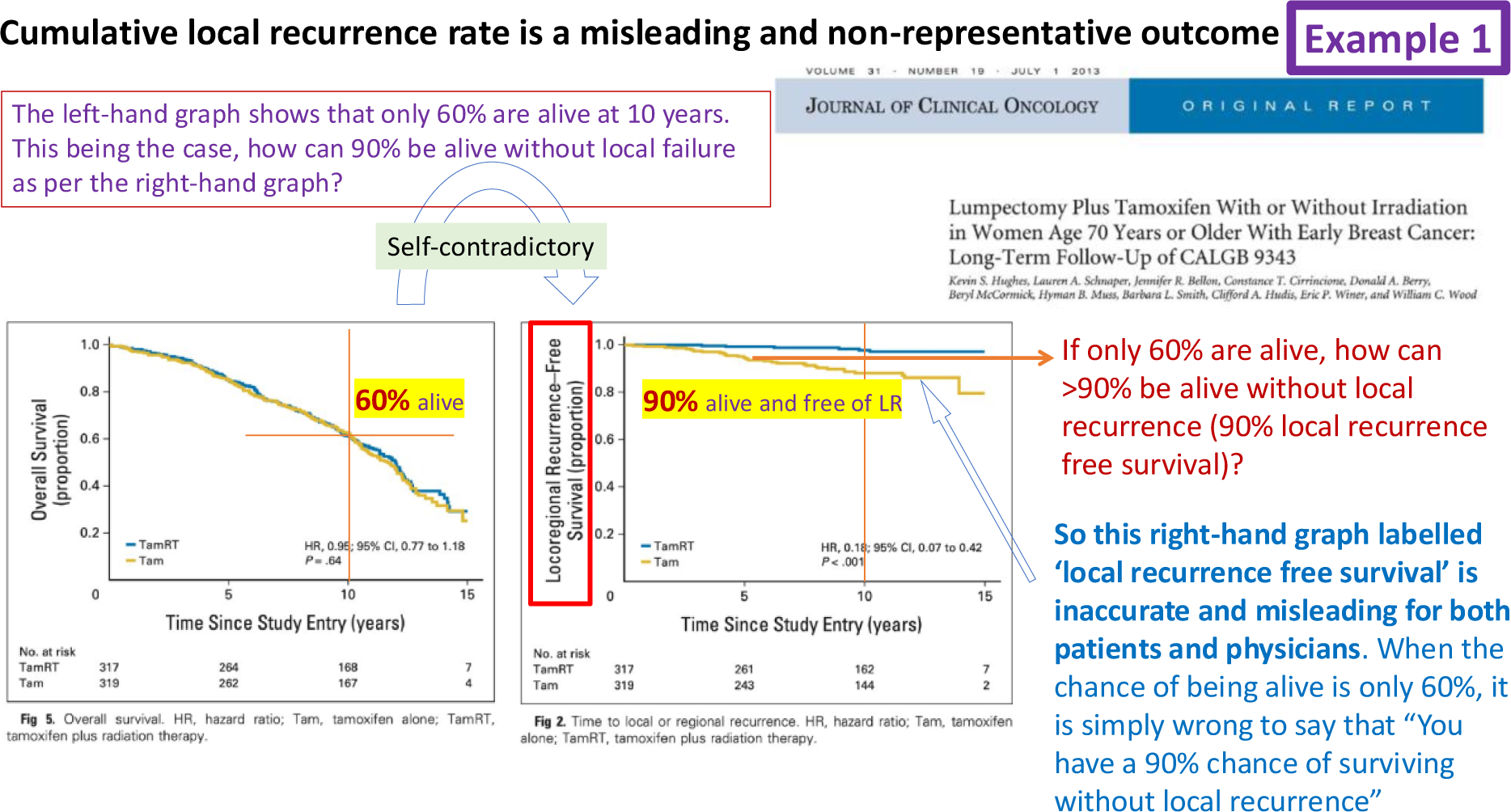
CALGB9343 trial: Radiotherapy vs no radiotherapy after lumpectomy.

**Example 2:**
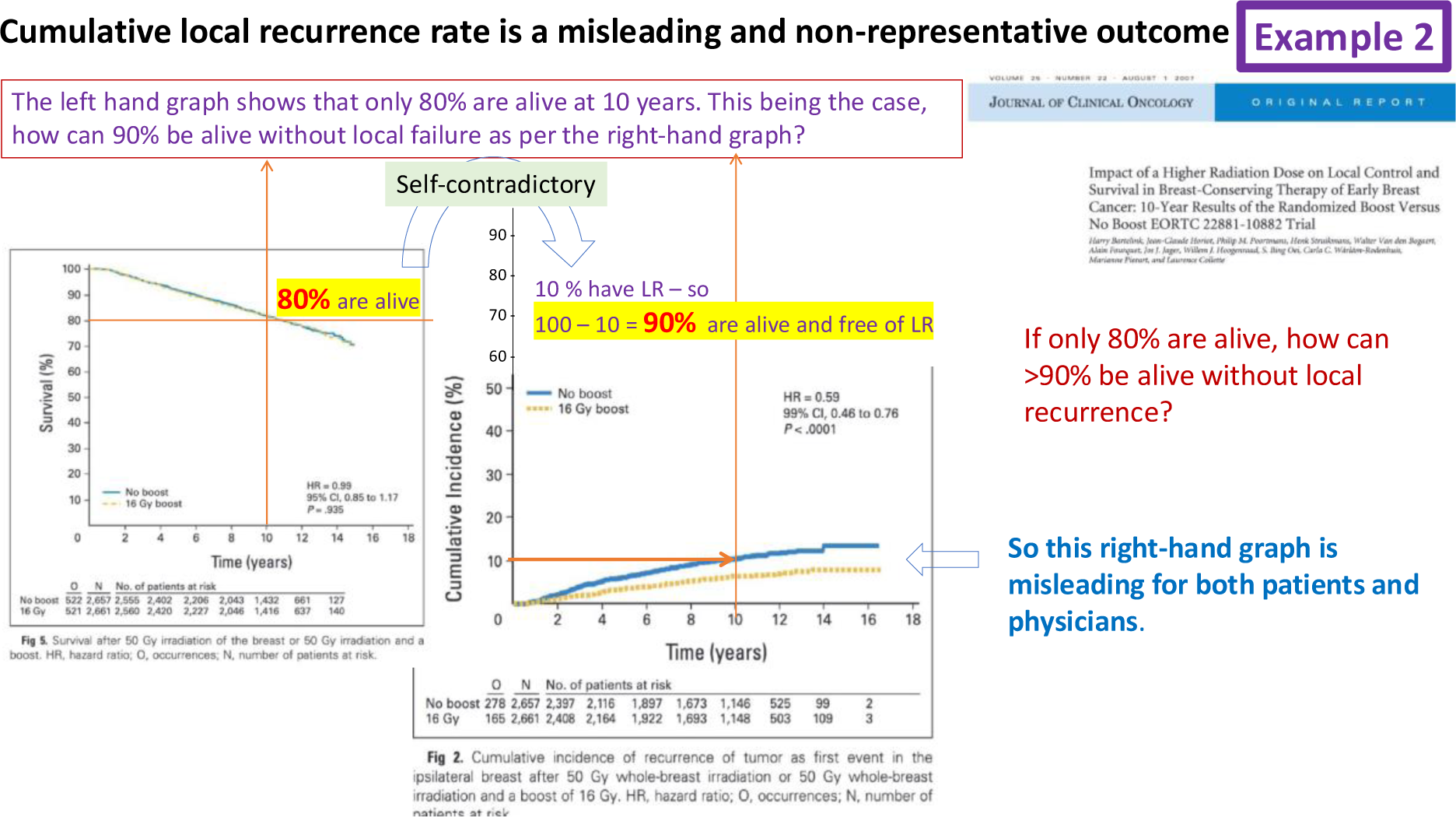
EORTC22881-10882 trial of tumour bed boost vs no boost.

**Example 3:**
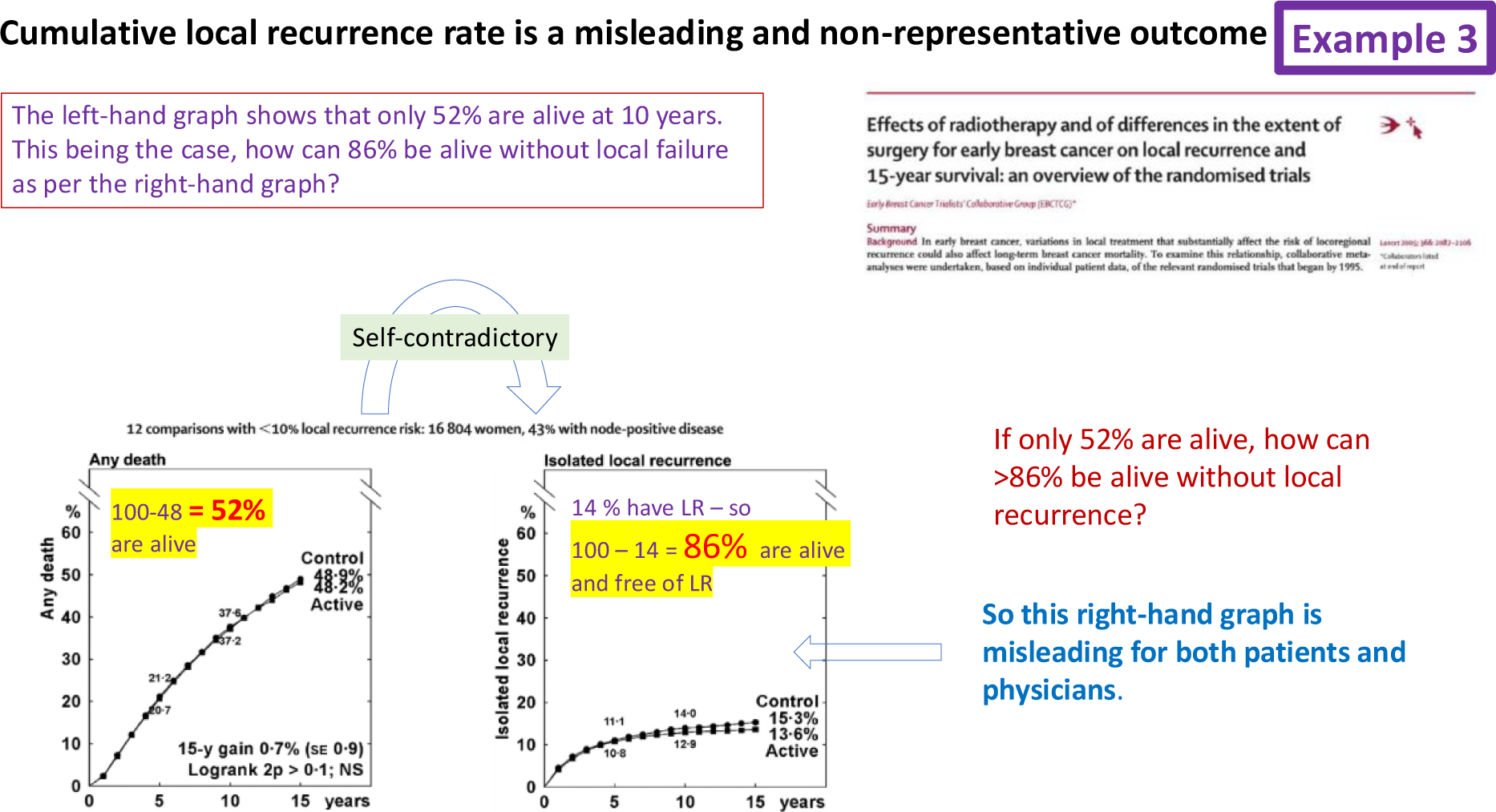
The Oxford overview meta-analysis of trials of radiotherapy vs no radiotherapy.

**Example 4:**
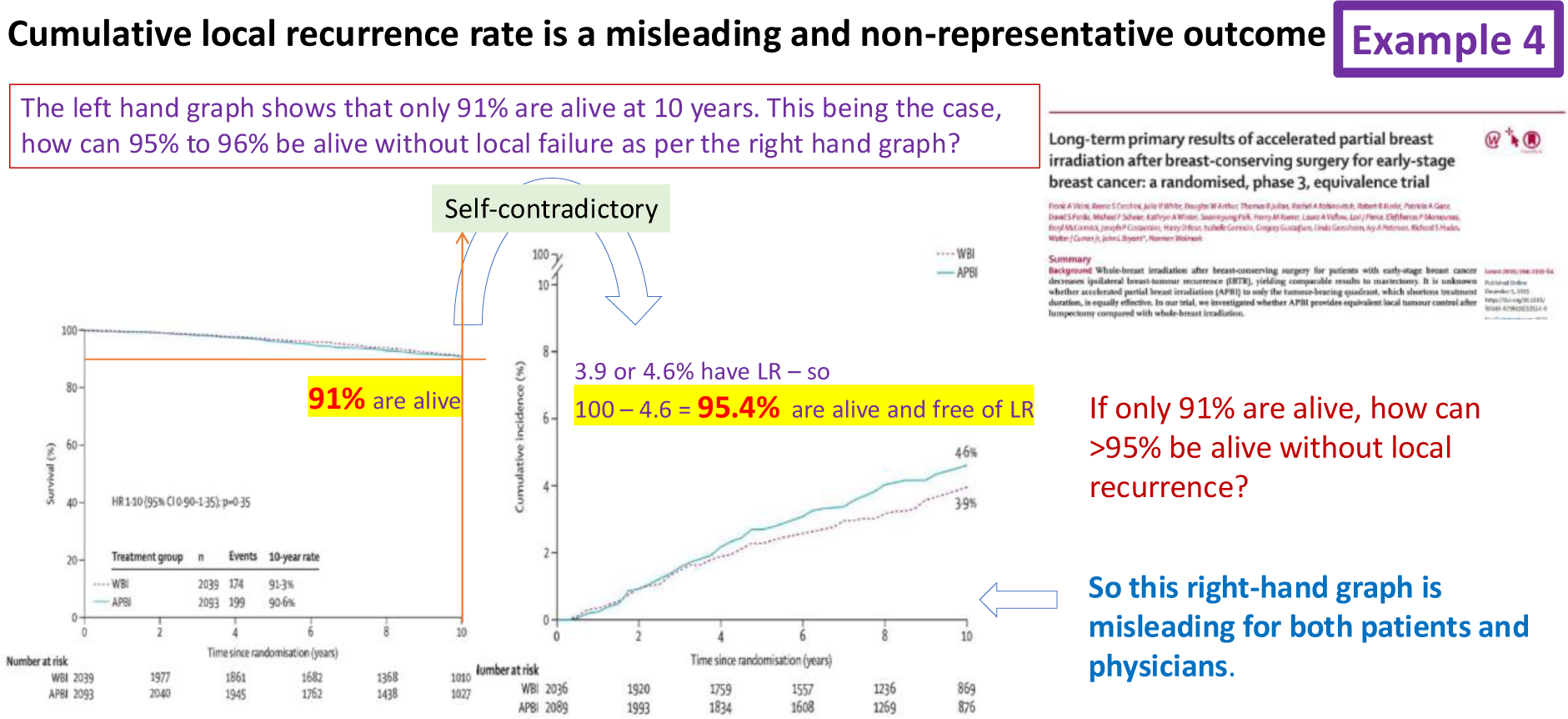
The NSABP-B39 trial of some methods of partial vs whole breast radiotherapy.

**Example 5:**
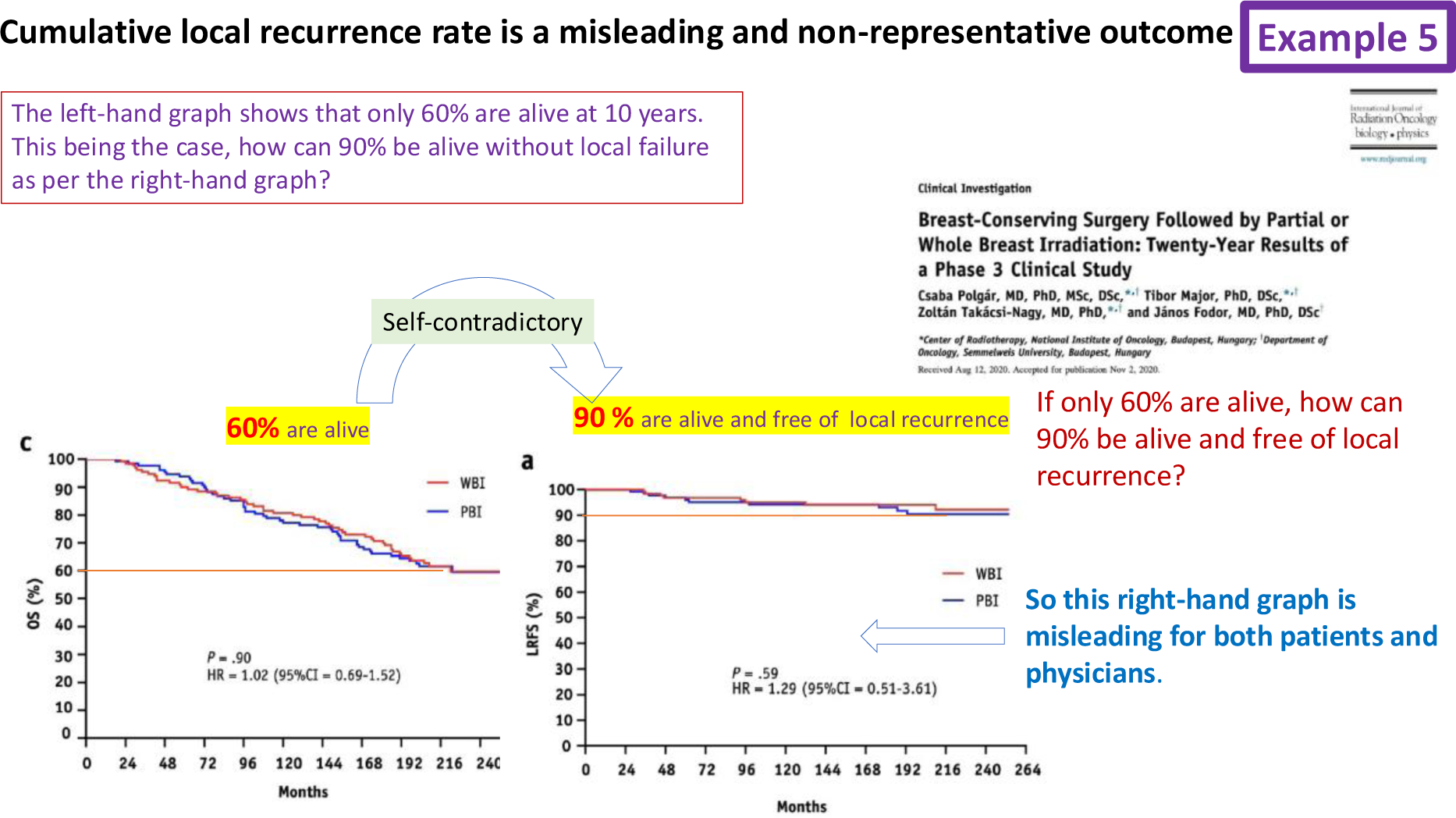
The Budapest trial of partial vs whole breast radiotherapy.

**Example 6:**
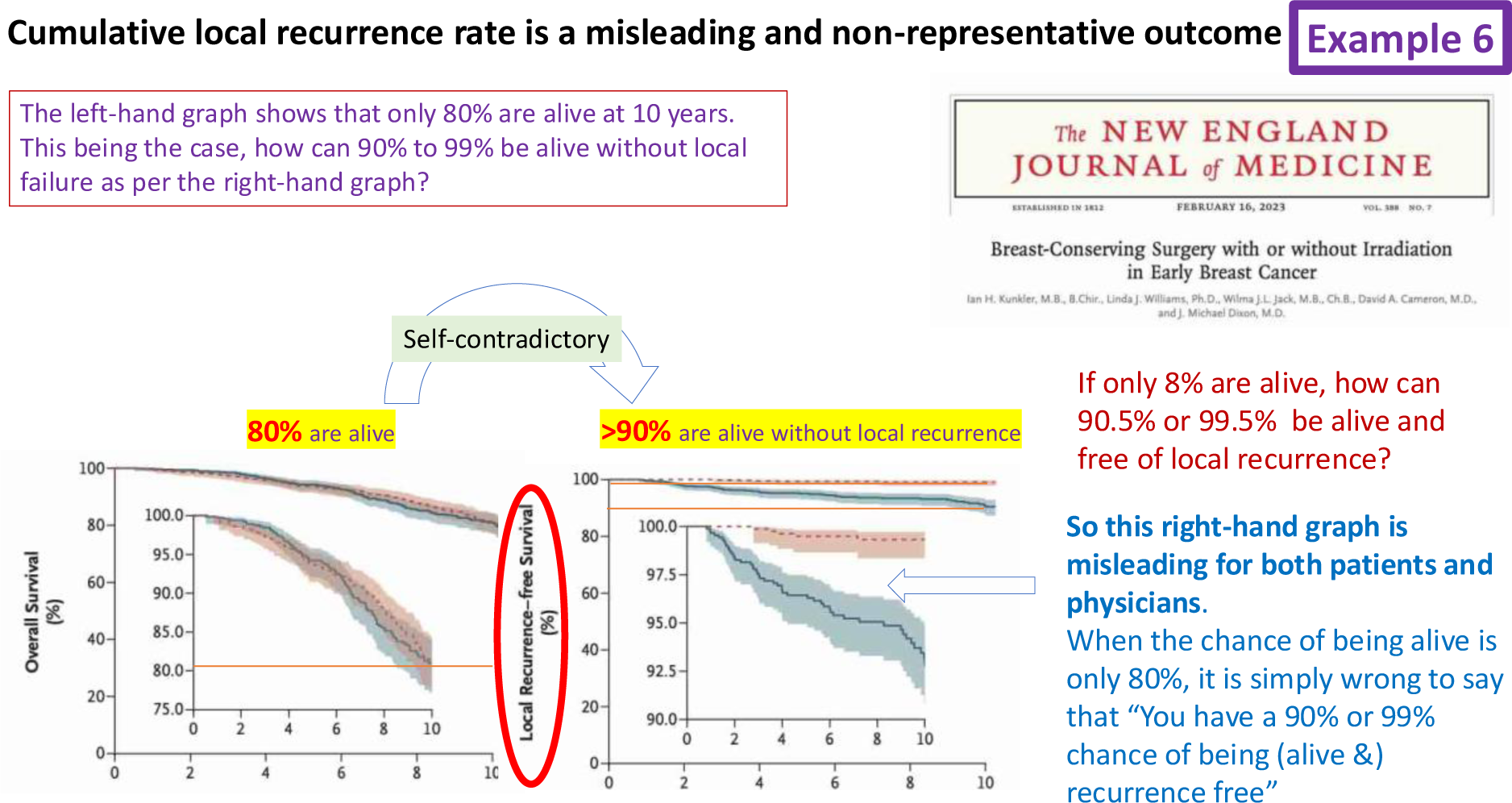
The PRIME-II trial of radiotherapy vs no radiotherapy in older /low risk women.

After the first six such examples, we present two examples where the error is avoided and the graphs illustrate what patients can truly show what happened to the patients in the trial and what they can reasonably expect to happen to them in the future if they choose to have the treatment(s).

**Table 1.** Summary of data shown in figures 1 to 6, showing how the local recurrence rates plotted by censoring the dead can result in misleading and non-representative figures.

| Example | Alive from OS graph | Alive from LR graph | Overestimate of % living - Absolute % | Overestimate of % living - Relative % |
| --- | --- | --- | --- | --- |
| 1: CALGB9343 trial | 60% | 90% | 30% | 150% |
| 2: EORTC22881-10882 trial | 80% | 90% | 10% | 112.5% |
| 3: The Oxford overview | 52% | 86% | 34% | 165% |
| 4: The NSABP-B39 trial | 91% | 95.4% | 4.4% | 105% |
| 5: The Budapest trial | 60% | 90% | 30% | 150% |
| 6: The PRIME-II trial | 80% | >90% | >10% | 112.5% |

### The recommended approach

In the two illustrations below, patients who had died were not censored leading to a more realistic representation of what patients can be expected to experience.

*Example 1:* In the TARGIT-A trial publications^11 12^, the Kaplan Meier curves comparing TARGIT-IORT vs EBRT take into account the fact that the patients who have died no longer carry a risk of local recurrence. Here the censoring is neither informative nor imbalanced. In these graphs, there is no contradiction between the overall survival and local-recurrence-free survival figures.

*Example 2*: Similarly, in the BIG 3-07/TROG 07.01 trial^14^, published after the TARGIT-A papers were published, the local control is expressed correctly so that the proportion of patients who are alive and free of local recurrence is less than the overall survival

N.B. Subtracting overall survival from local recurrence figures can overestimate the difference in ‘pure’ local recurrence, for example in the TARGIT-A trial, 5 times more patients died after local recurrence after EBRT (43% at 12 years) compared with those with local recurrence after TARGIT-IORT (9%)^12^.

### The recommended way

**Example 1.**
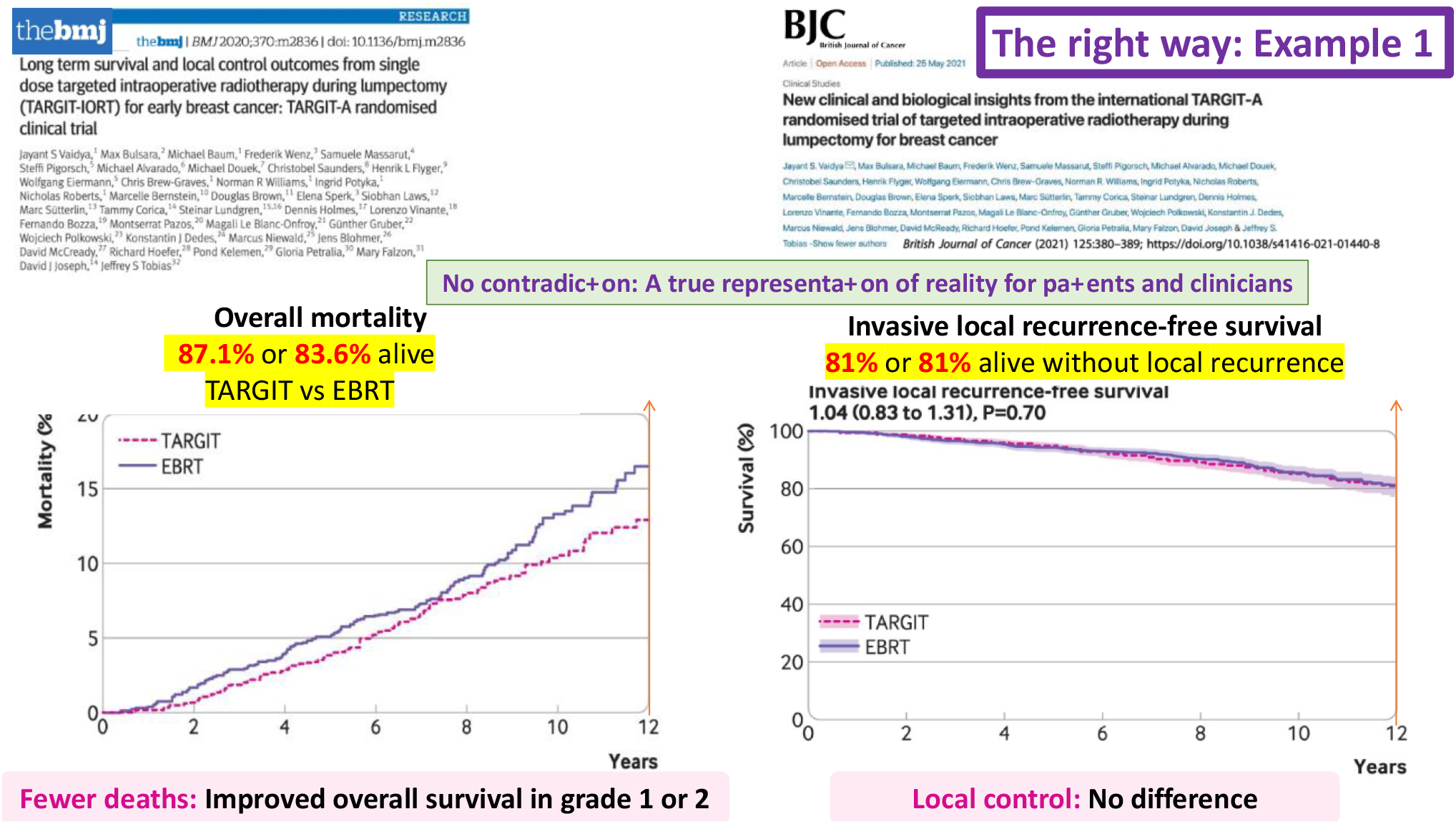
The TARGIT-A trial of targeted intraoperative radiotherapy vs whole breast radiotherapy.

**Example 2.**
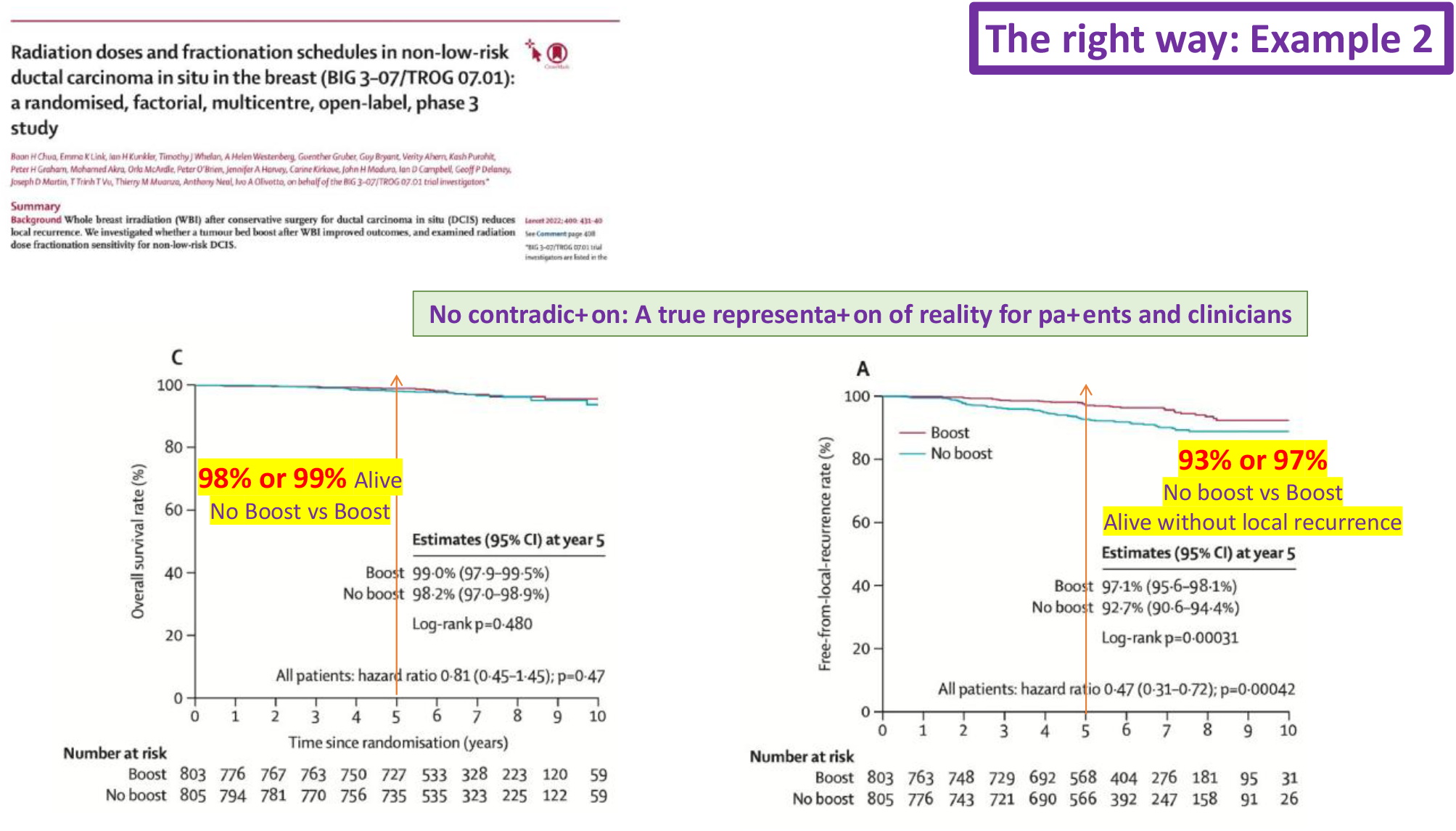
BIG 3-07/TROG 07.01 trial of tumour bed radiotherapy boost vs no boost.

## Discussion

Treatment of early breast cancer has evolved considerably over the past 100 years, and those diagnosed with breast cancer now have a very good life expectancy. Treatment related deaths now account for much larger proportion of deaths so counting all deaths while reporting the effect of any local treatment is increasingly important and the purist’s idea that they should not be counted is no longer valid. This principle is enshrined in IGH-GCP guidelines which states the use of “…end points that are well-defined, measurable, clinically meaningful and relevant to patients.”^i^ ^ii^The anomaly in the three examples of ‘cumulative incidence of local recurrence’ (examples 2, 3 and 4) or three of ‘local recurrence free survival’ (examples 1, 5 and 6) graphs is obvious and demonstrates the paradox that people are expected to have/not have a local recurrence when they are not even alive. This paradox occurs because deaths were *censored (considered alive)* while plotting these graphs. The most likely reason for employing such an obviously wrong approach is likely to be because of the (now disproven) preconception that radiotherapy affects only local breast tissues with no effect on mortality.

Furthermore, by statistical definition, *censored* patients should continue to have a risk of the event occurring, which is not true for dead patients, so they should not be censored. Also, the censoring must be balanced and independent of the treatment. It has been well established that radiotherapy can impact on death, independently of its local effect (e.g., by damaging the heart or inducing new cancers) and therefore censoring the dead is not independent of treatment and can be imbalanced if the effect on local control is in the opposite direction of the effect on mortality.

There are statistical methods that correct for *competing risks* such as death^15,16^. However, they still assume that the competing risk is independent of the event of interest and independent of the treatment arm. Either or both are not true in some scenarios. For example, local recurrence can impact distant disease and death; and whole breast radiotherapy (vs radiotherapy targeted only to the tumour bed) irradiates nearby vital organs such as the heart and the lungs and consequently impacts on resultant deaths from cardiovascular disease or other cancers.

Furthermore, corrections for competing risks (e.g. using the Fine and Gray model^15^) make only a small absolute difference in the estimates, and the resultant graphs would still suffer from the same misrepresentation of reality for the clinician and patients as demonstrated in the above figures. Saleh et al ^16^ have found a 28.4% relative difference between the EBCTCG estimates of local recurrence and those computed considering competing risks. As seen in figure 3, the EBCTCG estimate of % alive without local recurrence is in the range of 85% and applying Saleh et al’s relative difference of 28.4% would only change by 4% (28.4% of 15%). Such a small absolute change would still not be able answer the paradox – how can 80-90% of patients be around (alive) to be without local recurrence when only 60% are actually alive?

Graphs that supposedly represent cumulative incidence of local recurrence without counting the dead (and censoring them as if they are alive) in a quest for a ‘pure’ measure can be misleading for both patients and physicians. They do not provide meaningful understanding of what really happens to patients and cannot reliably provide a truthful comparison between the two treatments. The idea that a pure local recurrence graph represents “this is what would happen if you are still alive at 10 years” does not give a meaningful understanding because one then needs to factor in the ***untold*** chance of being alive at 10 years. We need to recognise this substantial deficiency and anomaly in such graphs, and they should not be published in the future. The correct measure for comparison of effects of a local treatment is local-recurrence-free survival.

### Here we explain why local recurrence free survival is not a ‘composite’ end-point

For assessment of local treatments, the desired patient outcome is ‘alive and *having no-local-recurrence’*.

Therefore, ‘***event’*** must be defined as the ***opposite of*** ‘alive and *having no-local-recurrence’*.

By definition, to have a local recurrence one needs to be alive, *i*.*e*., *not-dead!*

Thus ***opposite of*** ‘alive and *having no-local-recurrence*’ is **the *opposite of*** ‘*not-dead AND having-no-local-recurrence’* **which is ‘***death OR local recurrence’ which is must be definition of event. This is the definition used in the two examples above (the recommended approach)* The approach that we recommend is consistent with the European (DATECAN) and American (STEEP) guidelines^17 18^ which provide standardized definitions of time to event endpoints for cancer trials. They recommend that for assessment of local treatments, the time to event endpoint should be local recurrence free survival which includes deaths, whatever the cause, as events.

When comparing Kaplan-Meier curves, one should be taking the whole curve into account, rather than point estimates. In these examples, we have used point estimates not just for simplicity but also because these are the ones frequently used by clinicians, albeit wrongly, to consider the options for themselves and for their patients. The point we are trying to make – the huge discrepancy in the representation of reality by the ‘pure’ local recurrence curves and the actual reality – remains valid whether we consider the areas under the curves or just the point estimate.

Patients with breast cancer have numerous options to consider when embarking on treatment, and informed shared decision-making is essential. Discussions about local recurrence rates may often be part of discussions between doctors and patients. As we have explained, the method of presenting data we recommend here is far more readily understood than are data based on censoring of deaths, where the percentage “recurrence-free” at a given point in time is not directly related to the number of living patients at that time.

To summarize, ultimately, there are only two meaningful outcomes in the treatment of breast cancer, length of life and quality of life (QOL). Measuring length of life is self-evident but QOL requires complex “psychometric instruments”. A potential surrogate for QOL is avoidance of local recurrence, and its downstream consequences such as a mastectomy. The convention of describing this outcome has been cumulative local recurrence rate, but this is biased because the interventions that impact local recurrence also can independently impact length of life and quality of life and this impact may be larger in magnitude than the impact on local recurrence. The simple way to avoid this error is to turn the analysis on its head by reporting chance of success rather than failure by reporting chance of being free of local recurrence over a set period of time (including both death and local recurrence as events).

## Conclusion

In trials assessing local treatments, success – i.e., achieving local control (or preventing local recurrence), - is remaining alive without local recurrence, which is correctly measured by use of local recurrence free survival, in which both local recurrence and deaths are counted as events. This estimate truly represents what really happens to patients in terms of local control / local failure and the relative effectiveness of treatment(s). It also matches how ‘disease-free survival’ is estimated, as well conforming to the recommendations of European (DATECAN) and American (STEEP) guidelines^17 18^.

## Data Availability

All data produced in the present work are contained in the manuscript

## Acknowledgments

The authors thank Prof Michael Baum, Professor Emeritus of Surgery and Visiting Professor of Medical Humanities, University College London for his insightful comments and suggestions.

## Footnotes

i https://www.ema.europa.eu/en/documents/regulatory-procedural-guideline/ich-guideline-e8-r1-general-considerations-clinical-studies_en.pdf (section 3.1)

ii https://www.ema.europa.eu/en/documents/scientific-guideline/ich-e9-r1-addendum-estimands-and-sensitivity-analysis-clinical-trials-guideline-statistical-principles-clinical-trials-step-5_en.pdf

## Notes

### Competing Interest Statement

The authors have declared no competing interest.

### Funding Statement

This study did not receive any funding

